# Upregulation of Human Endogenous Retrovirus-K (HML-2) mRNAs in hepatoblastoma: Identification of potential new immunotherapeutic targets and biomarkers

**DOI:** 10.1101/2020.02.28.20026260

**Authors:** David F Grabski, Aakrosh Ratan, Laurie R Gray, Stefan Bekiranov, David Rekosh, Marie-Louise Hammarskjold, Sara K Rasmussen

**Affiliations:** Department of Surgery, University of Virginia School of Medicine; Myles H. Thaler Center for AIDS and Human Retrovirus Research, University of Virginia; Center for Public Health Genomics, Department of Public Health Sciences, University of Virginia School of Medicine; Department of Microbiology, Immunology and Cancer Biology, University of Virginia School of Medicine; Department of Biochemistry and Molecular Genetics, University of Virginia School of Medicine; Division of Pediatric Surgery, Department of Surgery, University of Virginia School of Medicine

**Keywords:** Human Endogenous Retrovirus-K, Hepatoblastoma, Fetal Tumor, Immunotherapy, Tumor Biomarker

## Abstract

**Purpose:** Hepatoblastoma is the most common liver malignancy in children. In order to advance therapy against hepatoblastoma, novel immunologic targets and biomarkers are needed. Our purpose in this investigation is to examine hepatoblastoma transcriptomes for the expression of a class of genomic elements known as Human Endogenous Retrovirus (HERVs). HERVs are abundant in the human genome and are biologically active elements that have been associated with multiple malignancies and proposed as immunologic targets in a subset of tumors. A sub-family of HERVs, HERV-K (HML-2), have been shown to be tightly regulated in fetal development, making investigation of these elements in pediatric tumors paramount.

**Methods:** We first created a HERVK-FASTA file utilizing 91 previously described HML-2 proviruses. We then concatenated the file onto the GRCh38.95 cDNA library from Ensembl. We used this computational tool to evaluate existing RNA-seq data from 10 hepatoblastoma tumors and 3 normal liver controls (GEO accession ID: GSE89775). Quantification and differential proviral expression analysis between hepatoblastoma and normal liver controls was performed using the pseudo-alignment program Salmon and DESeq2, respectively.

**Results:** HERV-K mRNA was expressed in hepatoblastoma from multiple proviral loci. All HERV-K proviral loci were expressed at higher levels in hepatoblastoma compared to normal liver controls. Five HERV-K proviruses (1q21.3, 3q27.2, 7q22.2, 12q24.33 and 17p13.1) were significantly differentially expressed (p-adjusted value < 0.05, |log2 fold change| > 1.5) across conditions. The provirus at 17p13.1 had an approximately 300-fold increased expression in hepatoblastoma as compared to normal liver. This was in part due to the near absence of HERV-K mRNA at the 17p13.1 locus in fully differentiated liver samples.

**Conclusions:** Our investigation demonstrates that HERV-K is expressed from multiple loci in hepatoblastoma and that the expression is increased from several proviruses as compared to normal liver controls. Our results suggest that HERV-K mRNA expression may find use as a biomarker in hepatoblastoma, given the large differential expression profiles in hepatoblastoma, with very low mRNA levels in liver control samples.

## 1. Introduction

Hepatoblastoma is the most common pediatric liver malignancy, affecting approximately 500 children in the US each year [1, 2]. Similar to other fetal tumors, hepatoblastoma is thought to arise from embryonic liver progenitor cells that fail to differentiate into hepatocytes [3-5]. As hepatoblastoma precursor cells show different levels of differentiation prior to malignant transformation, this cancer is morphologically complex and histologically subcategorized as one of the following subtypes: fetal, embryonal, or mixed epithelial and mesenchymal undifferentiated small cell [6]. Treatment is multimodal, involving a combination of resection and chemotherapy or transplant [7]. Five-year survival in North America is between 70-80%, with the best outcomes in early stage disease [8, 9]. There is still a clear need to identify novel treatment strategies that can offer more children hope for a long term cure [10, 11]. Furthermore, a full understanding of the molecular drivers of these tumors will be advantageous in the search for new treatments [12].

This report focuses on the expression of Human Endogenous Retrovirus-K (HERV-K) (HML-2) mRNA in hepatoblastoma and identifies HERV-K expression as a potential disease marker in this cancer. HERVs have garnered increasing attention in translational investigative science in the last 30 years as evidence has accumulated that these genomic elements have significant effects on human biology, both in health and disease. Attention has recently turned to the potential role of HERVs as both targets for immunotherapy [13, 14] as well as tumor markers to stratify disease in different solid organ malignancies [15, 16].

Human endogenous retroviruses (HERVs) are transposable genomic elements that have integrated into the human germline over many millions of years. All together, HERVs comprise an estimated 8% of the human genome [17]. Several HERVs have been co-opted by the human cell for specific biologic functions. For example, the Syncytin protein, responsible for the formation of syncytiotrophoblasts and ultimately mammalian placentation, was exapted from a HERV envelope protein [18]. In addition, a HERV viral promoter activates and drives transcription of the amylase gene in the hominid parotid gland, which may have been key to the expansion of the hominid diet to include starches [19]. For in-depth reviews of HERV biology, see [20-22].

There have been numerous phylogenetically distinct integrations of HERVs over evolutionary time with approximately 40 independently identified sub-groups [23]. The HERV-K (HML-2) viruses represent the most recent retroviruses to integrate into the human germline and have thus undergone the least amount of genetic silencing. In addition, these viruses remain polymorphic in human populations [24]. When transcriptionally active, some of the HERV-K proviruses are still capable of producing viral proteins, which can exert varied functions, such as nucleocytoplasmic export of mRNA and cell-cell fusion. There are approximately 90 identified intact or partially intact HERV-K (HML-2) proviruses distributed throughout the human genome, as well as over 900 solo long terminal repeats (LTRs) that are remnants of inserted viruses [25].

In the majority of human cell types and cell states, HERV-Ks have been transcriptionally silenced through DNA methylation and chromatin remodeling [26]. However, growing evidence suggests that there are multiple diseases and developmental states where HERV-K mRNAs are more highly expressed than in their normal adult somatic cell counter-parts [27-29]. Important to this investigation, HERV-K has been shown to be over-expressed in multiple tumors [30, 31], as well as during fetal development [32, 33], leading to increasing research into the potential effects of HERV-K expression in both oncogenesis and during embryogenesis.

In a previous study, Zhao et al. identified HERV-K envelope mRNA expression in breast cancer tissue, while expression was not detectable in healthy breast tissue controls [30]. Additional studies noted that HERV-K mRNA expression levels directly correlated with the eventual development of distant metastasis [15], while another investigation found that the overexpression of HERV-K was specifically associated with the basal cell subtype [34]. Similarly, in melanoma, HERV-K mRNA expression effectively distinguished tumor samples as well as metastatic lymph nodes from normal tissue and non-metastatic lymph nodes [31]. In an investigation of hepatocellular carcinoma and surrounding normal tissue it was noted that HERV-K mRNA levels were increased in hepatocellular carcinoma and that this provided an independent prognostic indicator for lower overall survival [35]. For the current understanding of HERV biology in cancer, see [36, 37].

Given the large differential expression profiles between disease and normal tissue and that HERV-Ks can express viral proteins capable of activating both innate, humoral and cell-mediated immune responses [38-42], attention has recently turned to the possibility of utilizing HERV-K neoantigens for targeted therapy. Chimeric antigen receptor (CAR) T cells that target HERV-K Env proteins have been developed and tested in *in vivo* murine models for both breast cancer and melanoma [43, 44]. In both models, the HERV-K Env CAR T-cells demonstrated tumor specific cytotoxicity, reduced the primary tumor mass and showed reduction of tumor metastases.

Importantly, HERV-K expression has remained unexamined in pediatric tumors. This oversight is potentially impactful, given that HERV-K has also been shown to be regulated and transcribed during embryogenesis and progressively silenced as fetal development continues [32, 33]. Transcriptional activity of HERV-K proviruses in pediatric tumors are thus of specific interest as these cancers are thought to arise from embryonic precursor cells that fail to differentiate during organ development. Given the increased expression of HERV-K mRNA in hepatocellular carcinoma, as well as the fact that hepatoblastoma fits the paradigm of a fetal tumor which arises from a failure of cellular differentiation, we sought to investigate HERV-K expression in this tumor. We hypothesized that HERV-Ks may be more expressed in hepatoblastoma, as compared to normal, fully differentiated liver tissue. In this investigation, we utilized publicly available RNA sequencing data from hepatoblastoma and normal liver controls generated by the Children’s Hospital of Pittsburgh [45]. We utilized standard bioinformatics techniques to create a custom database that enabled us to examine RNA expression from HERV-K proviruses in these data sets. Here, we report on differences in HERV-K RNA expression in hepatoblastoma compared to normal liver controls.

## 2. Methods

### 2.1 Hepatoblastoma and Normal Liver RNA-seq Data

The dataset used in this investigation includes RNA sequencing (RNA-seq) data from 10 hepatoblastoma samples and 3 normal liver controls. Tumor excision and RNA isolation was performed by the University of Pittsburg Children’s Hospital as part of a next-generation sequencing (NGS) study to identify activated cancer pathways in clinically aggressive hepatoblastoma [45]. The raw sequencing data are publicly available and were downloaded from the NCBI biorepository using the NCBI Sequence Read Archive (SRA) Toolkit (GEO accession ID GSE89775). According to the NCBI biorepository, total RNA (1 ug) was isolated from fresh frozen tissue (both hepatoblastoma and normal liver) and sequenced on an Illumina platform to generate 100 base-pair, strand-specific, paired-end reads to a sequencing depth of approximately 40M reads per sample. Prior to analysis in this study, all raw FASTQ files were pre-processed with Trimmomatic to remove adaptors and low-quality reads as well as to assure that only paired-ends reads with a minimum read length of 50 nucleotides were included [46]. The quality of the raw and trimmed reads from each sample was confirmed with the program FASTQC (http://www.bioinformatics.bbsrc.ac.uk/projects/fastqc).

### 2.2 Analysis of HERV-K mRNA expression

HERV-K proviruses are currently not well annotated in the human genome, meaning that standard RNA-seq analysis techniques cannot be used to determine the expression profile of HERV-Ks. To overcome this limitation, we created a HERV-K specific FASTA file using the genomic sequence of the 91 HERV-K proviruses deposited in NCBI (GenBank ID JN675007-JN675097). To determine the transcriptional profile and differential expression of HERV-K in all samples, we concatenated our working HERV-K FASTA file onto the GRCh38.95 cDNA fasta file downloaded from Ensembl. For RNA analysis, we did not annotate the individual potential spliced transcripts that would be expected to be expressed from an integrated provirus, but rather defined the entire proviral sequence as a single transcript. We then used the pseudo-aligner, Salmon [47] in mapping-based mode with the validateMappings flag to create a count matrix over the full human transcriptome including the concatenated HERV-K file (example code: salmon quant -i GRCh38_HERVK.fa -l -1 FT6_1.fq -2 FT6_2.fq –validateMappings -o FT6_quant). We then sub-selected read counts assigned to HERV-K loci.

We next indexed the HERV-K FASTA file with the alignment tool HISAT2 [48] and aligned our Hepatoblastoma and Normal Liver samples to the HISAT2-HERV-K index to create .BAM files. To control for multi-mapping of the repeat elements, we used SAMTOOLS to select for reads with a mapping quality (MAPQ) Score >= 50, which has been shown to be an effective way to eliminate multi-mapped reads from HERV-K analysis [49]. Uniquely aligned .BAM files for all samples were then imported into the software package Geneious (Biomatters, Auckland, New Zealand) for visualization of read position along an annotated HERV-K provirus which distinguishes reading frames for the different viral proteins.

### 2.3 HERV-K Expression Profiles and Differential Expression

Transcript abundance read estimates from Salmon were imported into R (version 3.5.1) using tximport [50]. Transcript abundance estimates were normalized for sample sequencing depth using the R Bioconductor package DESeq2 [51]. This allowed us to determine normalized HERV-K expression across all proviral loci by sample, together with the number of loci responsible for total HERV-K expression and the range of reads across each locus. Differential expression of HERV-K in hepatoblastoma as compared to normal liver was also analyzed using DESeq2. HERV-K proviruses were considered differentially expressed if the p-adjusted values (calculated using the Benjamini-Hochberg False Discovery Rate implemented in DESEq2) were less than 0.05 and the absolute value of the log2 fold changes were greater than 1.5 [52].

Given the apparent heterogeneity in HERV-K expression across different hepatoblastoma samples, we also stratified hepatoblastoma samples by overall HERV-K expression (total number of normalized reads across all proviral loci). We selected the top 3 highest HERV-K expressing hepatoblastoma samples and the three lowest HERV-K expressing hepatoblastoma samples. We then performed a differential gene expression analysis again using DESeq2 to compare the high expressing to low expressing tumors. Genes with a p-adj value < 0.05 and |log2 fold change| > 1.5 were considered significant and included in a Gene Ontology (GO) and Kyoto Encyclopedia of Genes and Genomes (KEGG) over-representation functional analysis. Gene pathways were considered enriched if they had a p-adj value < 0.05. GO and KEGG analysis was performed using the clusterProfiler package in R [53].

Unless otherwise specified, all plots denoting the RNA expression profile and differential expression were generated using the ggplot2 package in R [54]. The scatterplot was created using the EnchancedVolcano visualization package in R [55].

## 3. Results

The RNA-seq that was downloaded from the NCBI biorepository included 10 hepatoblastoma samples (HB) from children undergoing liver transplantation for their disease. These samples represent aggressive hepatoblastomas that were not amenable to up-front resection. It also included 3 normal liver controls (NC) from orthotopic livers prior to transplant. Following pseudoalignment with Salmon and sample normalization with DESeq2 as described in the Methods, we found that the HERV-K RNA expression profile varied greatly across the hepatoblastoma (HB) samples and normal liver controls (NC). In HB, the median HERV-K read counts across all proviral locations for each sample was 342 (interquartile range (IQR) 235, 515). However, 2 samples had greater than 2,000 reads that aligned to several HERV-K proviruses, whereas 2 samples showed 150 read counts or less (Table 1). The general HERV-K expression profile across all samples is visualized in the HERV-K expression heatmap in Figure 1. Each cell in the heatmap represents the normalized expression Z-score calculated across samples (range -3 to 3) of a specific HERV-K provirus (y-axis) in individual HB or NC samples (x-axis). mRNA from numerous proviral loci were expressed at average levels across all the samples (Z-score = 0), which we confirmed were HERV-Ks expressed at low levels (average of less than 10 reads across a HERV-K provirus in individual samples). These HERV-K loci are represented by light blue in the heatmap. There was great heterogeneity in proviral expression in individual samples, including variation among the hepatoblastoma samples (9.5 expressed proviral loci, IQR 9.3). Variation was also was observed in the normal liver controls (3 expressed proviral loci, IQR 3) although the total number of expressed HERV-K loci was less than in HB. No proviral loci were common to all samples and many HERV-K loci were expressed in less than 5 total samples.

**Table 1:** Normalized HERV-K read counts and number of transcribed HERV-K loci in hepatoblastoma and normal liver controls

| Sample | Normalized Read Count<br>across all HERV-K Loci | Total HERV-K locations<br>with > 10 normalized reads | Range of Reads Across<br>Individual Provirus |
| --- | --- | --- | --- |
| **Hepatoblastoma** |  |  |  |
| FT6 | 273.2 | 10 | 10.4- 29.5 |
| FT7 | 223.3 | 5 | 12.6- 74.6 |
| FT8 | 532.0 | 3 | 20.1- 363.8 |
| FT9 | 2,778.8 | 42 | 14.4- 243.5 |
| FT10 | 148.3 | 1 | 10.1 |
| FT11 | 2,503.8 | 28 | 22.2- 410.3 |
| FT12 | 270.4 | 9 | 10.4- 26.9 |
| FT13 | 465.1 | 13 | 10.5- 162.1 |
| FT14 | 412.2 | 12 | 10.2- 51.2 |
| FT15 | 153.0 | 2 | 11.9-35.1 |
| **Normal Liver** |  |  |  |
| NC1 | 141.2 | 3 | 11.5- 50.3 |
| NC2 | 98.1 | 1 | 14.6 |
| NC3 | 203.4 | 7 | 11.6- 23.4 |

Abbreviations: FT- Fetal Tumor (Hepatoblastoma), NC- Normal Control (Normal Liver)

**Figure 1:**
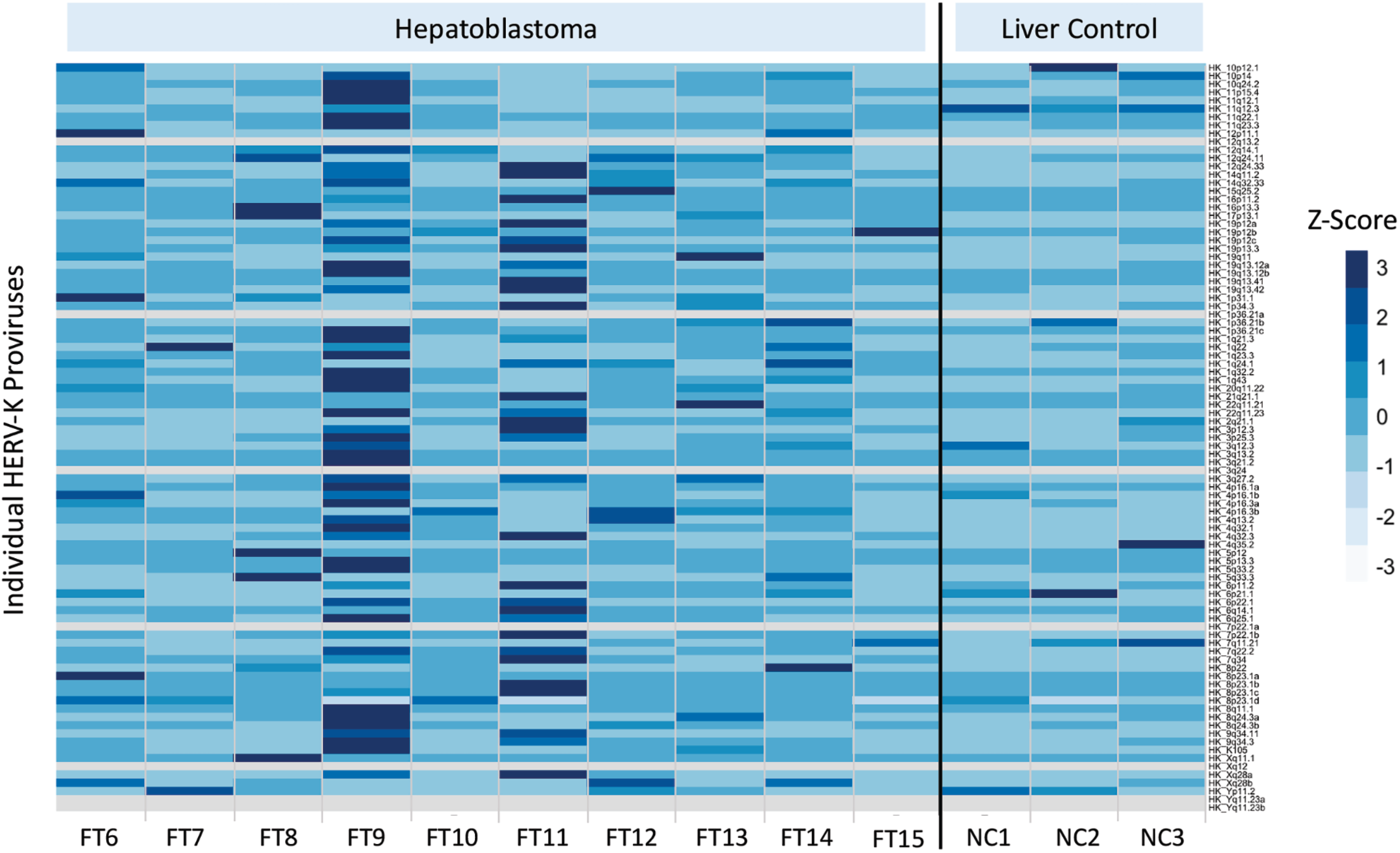
Heatmap of HERV-K expression in hepatoblastoma and normal liver controls. All tumor samples and normal liver controls are represented on the x-axis. All HERV-K proviral loci are represented on the y-axis. Color expression key is located in upper left of figure and is based on the calculated Z score (scale -3 to 3) across all samples in the study. Each individual cell represents the normalized HERV-K proviral expression (x-axis) in the respective sample (y-axis).

We next calculated differential proviral expression across the HB and NC samples. All proviral loci were expressed at higher levels in HB compared to NC as demonstrated by the HERV-K expression profile scatter plot (Figure 2). Five proviruses (1q21.3, 3q27.2, 7q22.2, 12q24.33 and 17p13.1) were significantly differentially expressed (p-adjusted value < 0.05, |log2 fold change| > 1.5) across conditions (Table 2). Boxplots of the log10 normalized counts for the HB and NC samples across each differentially expressed provirus reveal much higher expression in HB compared to NC samples (Figure 3). For 3 proviruses (17p13.1, 3q27.2 and 7q22.2), the normalized expression across all 3 NC samples was less than 10 reads and was too low for graphical comparison. The absence of expression in NC led to large fold-changes between HB and NC. Thus 17p13.1 showed a 294-fold increase expression in HB as compared to NC (padj = 0.009). This provirus was expressed in all hepatoblastoma samples with one exception and was not expressed in 2 of the 3 normal controls. Similarly, in HB samples, 7q22.2 was expressed 93.1-fold above NC (padj = 0.027), while 3q27.2 was expressed 55-fold above NC (padj = 0.026).

**Table 2:** Differential gene expression of HERV-K between hepatoblastoma and normal liver controls

| **HERVK Provirus** | **Log2 Fold Change** | **Log2 Fold Change**<br>**Standard Error** | **P-Value** | **P-adjusted Value** |
| --- | --- | --- | --- | --- |
| 1q21.3 | 4.0397 | 1.3917 | 0.00369 | 0.03891 |
| 3q27.2 | 5.8226 | 1.8781 | 0.00193 | 0.02645 |
| 7q22.2 | 6.5405 | 2.1196 | 0.00203 | 0.02713 |
| 12q24.33 | 5.3952 | 1.6643 | 0.00119 | 0.01941 |
| 17p13.1 | 8.2095 | 2.3046 | 0.00037 | 0.00927 |

**Figure 2:**
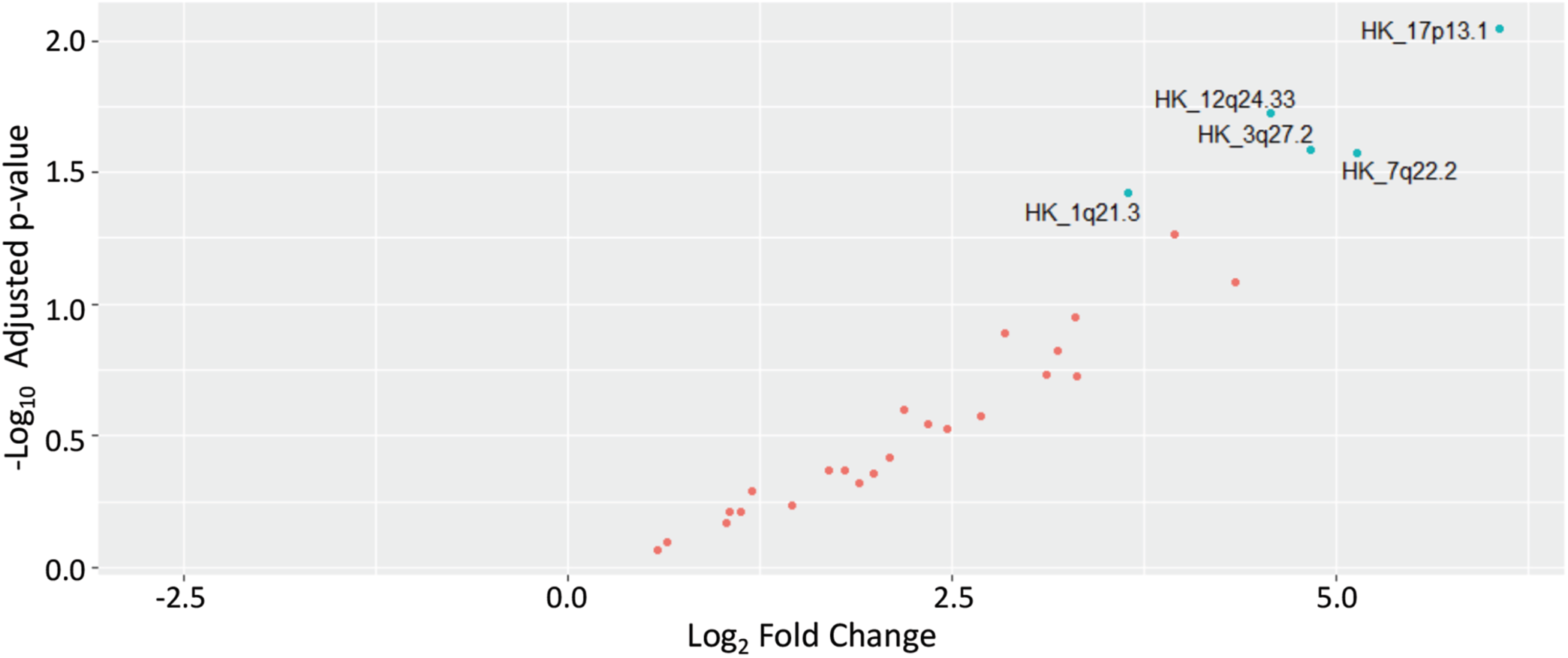
Scatter plot of HERV-K differential expression profile between hepatoblastoma and normal liver controls. Each point on the figure represents the log2 fold change (x-axis) between conditions of an individual HERV-K provirus plotted against the corresponding -log10 p-adjusted value (y-axis). Orange points represent HERV-K proviruses that were not significantly differentially expressed (p-adj < 0.05, |log2fold change| > 1.5) between conditions. Green points represent differentially expressed provirus, which are also labeled by genomic location (HK_1q21.3 represents the HERV-K provirus located at 1q21.3).

**Figure 3:**
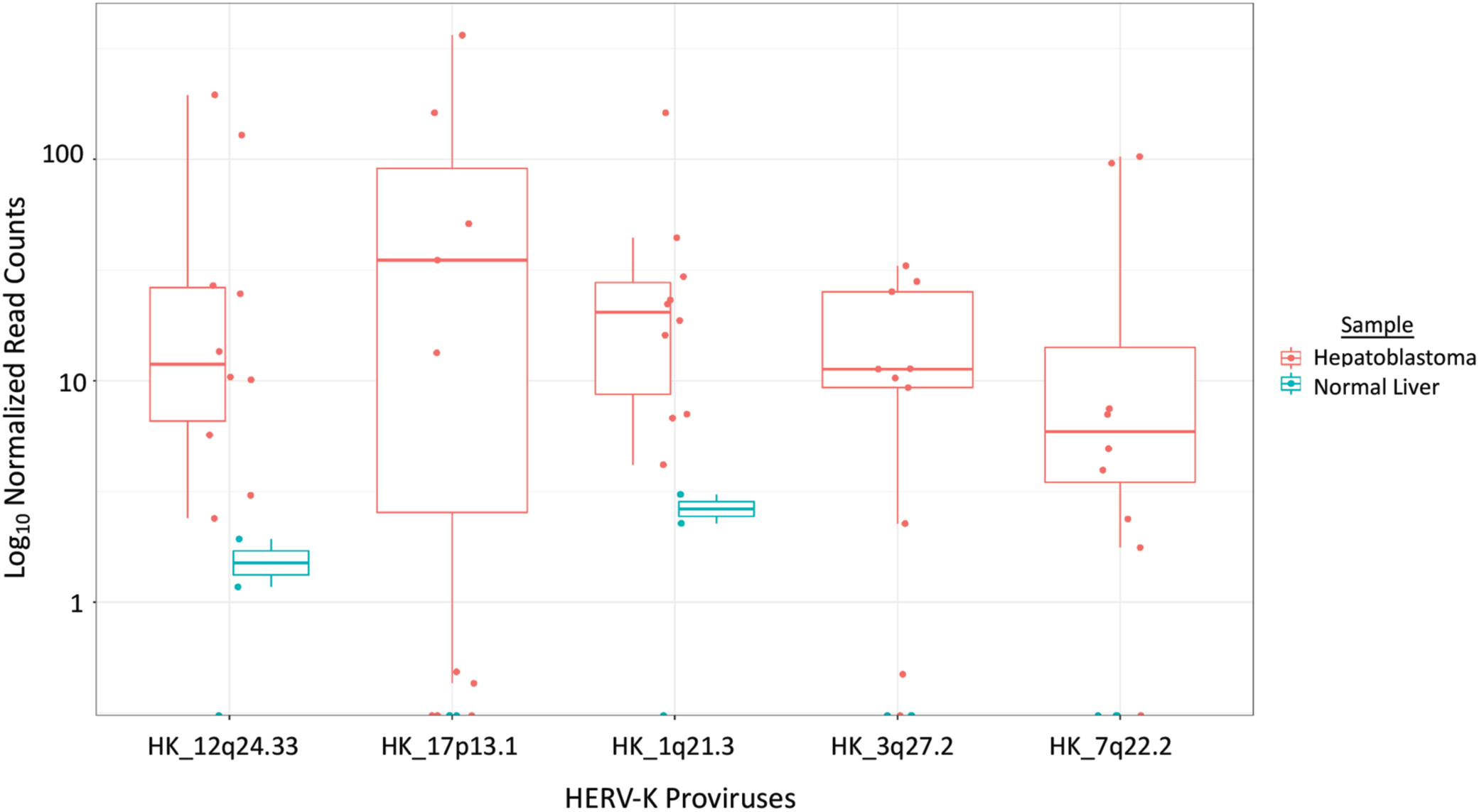
Boxplot and overlaid dotplot of significantly differentially expressed HERV-K proviruses. Individual proviruses are represented on the x-axis. Log10 normalized count values are represented on the y-axis. The normalized count value for each provirus in each sample is represented by an individual colored point. Hepatoblastoma samples are represented in orange while normal liver controls are represented in green. The central line of the boxplot is determined by the median log10 normalized expression value across all grouped samples (hepatoblastoma or normal liver controls) for each significantly differentially expressed provirus. The ‘box’ represents the 25^th^ (lower line) and 75^th^ (upper line) percentile log10 normalized expression across grouped samples.

Following alignment of all HB and NC samples with our HISAT2-HERV-K index and selection for uniquely mapped reads, we imported the alignment file (.BAM) into the bioinformatics platform Geneious. This allowed visualization of where the reads aligned across each provirus in the different samples. For provirus 17p13.1, the reads aligned across the entire provirus (Figure 4, panel A). Reads similarly mapped across the length of the provirus at 12q24.33 (Figure 4, panel B), with a small concentration of reads in the 5’ LTR (long terminal repeat). A similar pattern was seen in provirus 1q21.3, but with a concentration of reads in the 3’ LTR (Figure 4, panel C). Interestingly, despite 3q27.2 being a relatively complete provirus (9,100 bp), the majority of the reads aligned in the 3’ LTR for all samples (Figure 4, panel D). Conversely, all of the reads for 7q22.2 aligned uniquely to the 5’ LTR in all samples (Figure 4, panel E). Larger images for each individual provirus in Figure 4 are provided in Supplemental File 1.

**Figure 4:**
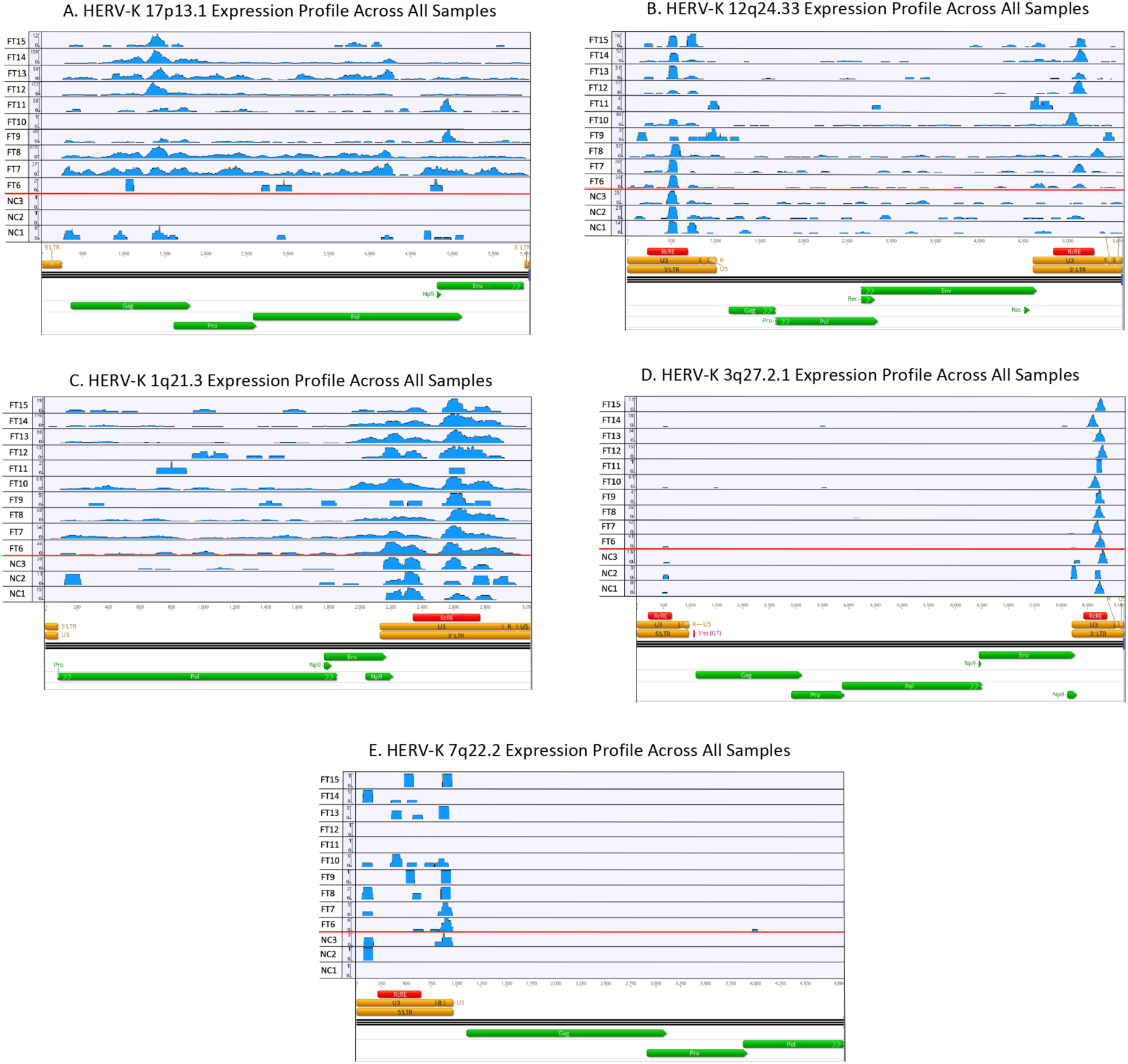
Graphical representation of uniquely aligned reads across HERV-K provirus (A)17p13.1 (B) 12q24.33 (C) 1q21.3 (D) 3q27.2 and (E) 7q22.2 created in bioinformatics platform Geneious. The x-axis represents the genomic position along the provirus. Major annotated regions of the proviral genome at each provirus are illustrated at the bottom of the panel. Coding regions for viral proteins Gag, Pro, Pol, Env, Rec or Np9 are represented by green bars, but does not necessarily infer an open-reading frame for the protein. Individual reads from each sample are represented on the y-axis. Abbreviations: FT-fetal tumor (hepatoblastoma), NC-normal control (liver).

Hepatoblastoma samples FT8 (531 HERV-K reads), FT9 (2,778 HERV-K reads and FT11 (2,503 HERV-K reads) represented the tumor samples with the highest HERV-K expression. Conversely, FT7 (223 HERV-K reads), FT10 (148 HERV-K reads) and FT15 (152 HERV-K reads) represented the tumors with the lowest expression. A differential expression analysis comparing the high to low tumors revealed 775 differential expressed genes (Supplemental File 2). GO Biological Process enrichment analysis of the differentially expressed genes revealed over-representation of cellular processes involved in leukocyte activation and leukocyte mediated immunity (Table 3). Additional GO terms (top 20) for cellular localization and molecular function as well as enriched KEGG terms are provided in Supplemental File 3.

**Table 3:** Gene Ontology Biological Process Enrichment Analysis following differential gene expression analysis of high HERV-K expressing HB vs low HERV-K expressing HB.

| **Functional Category** | **Differentially Expressed Genes** | **Total Genes in Functional Pathway** | **Enrichment False Discovery Rate (Adjusted p-value)** |
| --- | --- | --- | --- |
| Regulated exocytosis | 82 | 901 | 1.33E-12 |
| Exocytosis | 85 | 1023 | 2.41E-11 |
| Neutrophil activation | 61 | 594 | 2.41E-11 |
| Granulocyte activation | 61 | 603 | 2.55E-11 |
| Myeloid leukocyte activation | 69 | 766 | 8.96E-11 |
| Myeloid leukocyte mediated immunity | 62 | 647 | 8.96E-11 |
| Neutrophil mediated immunity | 59 | 591 | 8.96E-11 |
| Secretion | 124 | 1861 | 8.96E-11 |
| Myeloid cell activation involved in immune response | 61 | 640 | 1.60E-10 |
| Neutrophil activation involved in immune response | 57 | 583 | 3.11E-10 |
| Cell activation | 109 | 1591 | 4.16E-10 |
| Neutrophil degranulation | 56 | 577 | 5.59E-10 |
| Secretion by cell | 114 | 1715 | 6.51E-10 |
| Leukocyte degranulation | 59 | 632 | 6.51E-10 |
| Vesicle-mediated transport | 134 | 2220 | 5.32E-09 |
| Immune effector process | 96 | 1392 | 5.32E-09 |
| Leukocyte mediated immunity | 74 | 965 | 1.11E-08 |
| Leukocyte activation | 96 | 1416 | 1.21E-08 |
| Cell activation involved in immune response | 66 | 819 | 1.48E-08 |
| Leukocyte activation involved in immune response | 65 | 815 | 3.06E-08 |

\*Table represents the top 20 functional categories in the Gene Ontology Biological Process Enrichment Analysis stratified by false discovery rate.

## 4. Discussion

The data in this investigation establish that HERV-Ks are expressed in hepatoblastoma. Furthermore, the mRNA profile of HERV-K in hepatoblastoma is complex, with multiple proviruses transcribed from different loci in tumors from different individuals. The data also show that overall HERV-K expression is increased in hepatoblastoma compared to normal liver controls and that several proviruses show large fold increases in tumors compared to normal liver tissue. The significant increase makes HERV-Ks intriguing targets for immunotherapy. In addition, our data suggest that they could also serve as potential biomarkers for disease recurrence or progression, though further studies will be required to confirm this. This investigation is the first to demonstrate HERV-K expression in a pediatric solid organ malignancy. In addition, the bioinformatics pipeline described in this manuscript provides an effective tool to measure HERV-K RNA profiles in disease versus non-disease states that could be used to screen other fetal malignancies for HERV-K expression.

### 4.1 Expression of HERV-K in Hepatoblastoma and Potential Clinical Applications

Multiple HERV-K proviruses showed increased expression in hepatoblastoma. The HERV-K provirus at 17p13.1 was the most dramatic example of a large differential expression value, with an almost 300-fold change in expression from the provirus compared to normal liver tissue. Similarly, large differential expression values were seen for proviral loci 1p21.3, 3q27.2, 7q22.2 and 12q24.33.

The magnitude of the increased expression levels over normal liver controls was prominent in several instances. This is in part because mRNA levels from HERV-K proviruses in normal liver control were either not present at all, or present at very low levels (less than 10 read counts across the entire provirus). Our findings of low HERV-K expression in fully differentiated liver is consistent with previous investigations that have examined HERV-K expression in liver tissue [35, 56]. This finding is also consistent with the reported low levels of HERV-K expression in the majority of fully differentiated somatic tissues [35, 56].

Proviral expression profiles also differed across the hepatoblastomas themselves. Several hepatoblastoma samples had over 100 read counts aligned to the provirus at 17p13.1, while several other tumors had less than 10 counts (which was more similar to the expression profile in NC). This variation in proviral expression across HB samples was true for total HERV-K expression as well. Two hepatoblastoma samples had over 2,000 normalized read counts summed over all proviruses. In contrast, two HB samples had ∼150 normalized read counts across all proviruses with only 1 or 2 proviral locations with over 10 read counts. The HERV-K RNA expression in these tumors was thus more similar to the normal liver controls than to the other tumor samples. In follow-up investigations it will be important to determine whether HERV-K expression correlates with the molecular subtype of hepatoblastoma.

Furthermore, a differential gene expression analysis and GO enrichment analysis comparing high and low HERV-K expressing tumors demonstrated a strong correlation with cellular pathways involving leukocyte activation as well as neutrophil and leukocyte mediated immunity. Cellular pathway enrichment analyses such as GO and KEGG are useful adjuncts to understand how a complex set of differentially expressed genes may regulate specific biological process or known phenomenon. Our data suggest that HERV-K mRNA levels may correlate with either direct tumor immunogenicity or an inflammatory microenvironment surrounding the tumors. The possibility that expressed HERV-K proteins in these tumors could be acting as cancer neoantigens is an intriguing possibility, but this will require additional experimental validation. In a recent investigation, Rooney et al. analyzed approximately 20 solid organ tumors as well as normal tissue controls from TCGA mRNA-seq datasets for expression of both endogenous retrovirus (ERV) families as well as cytolytic activity. The investigation found that several tumor specific ERVs existed across multiple tumors and that high gene expression of these tumor specific ERVs correlated with significantly enriched immune activation pathways [57]. In addition, experimental evidence has demonstrated that HERV-K viral proteins can directly activate both innate and adaptive immune response in multiple solid organ tumors including breast cancer, melanoma and colorectal cancer [38-42].

The read distribution across individual proviruses varied but was consistent for each provirus across all samples in this study. The proviruses located at 17p13.1, 12q24.33 and 1q21.3 had read counts distributed across the entire provirus. 17p13.1 and 1q21.3 do not have intact 5’ LTRs suggesting that transcription is driven by an upstream cellular promoter and proviral expression represents read-through transcription from a cellular promoter. This phenomenon has been previously described [58]. Interestingly, the provirus at 3q27.2 had the majority of reads located in the 3’ LTR region, suggesting that transcription was initiating at the 3’ LTR. In 7q22.2, reads in all samples aligned to the 5’ LTR but ceased before progressing through the viral genome suggesting that either the 5’ polyA site was being utilized or epigenetic modifications silenced expression. Another important possibility is that this expression phenomenon of all reads aligning in the LTR (either 5’ or 3’) is associated with multi-mapped reads to an unannotated HERV-K LTR that is not included in our annotation file. While we controlled for multi-mapped reads in our bioinformatic pipeline, our analysis is dependent on ‘known’ HERV-Ks included in NCBI. This particular aspect of the analysis highlights the importance of detailed alignment. For example, 3q27.2 has an open reading frame for viral proteins Gag and Pol. However, in the case of this provirus, all the reads align in the 3’ LTR, and thus no proteins will be produced. It is important to note that transcription from the 5’LTR or 3’LTR of HERV-K proviruses may still be of biological significance as proviral enhancers can affect transcription of cellular genes up to 100,000 base pairs upstream or downstream of the HERV-K provirus [59].

Similar to the findings of our current investigation in Hepatoblastoma, increased HERV-K expression has been associated with multiple solid organ malignancies when compared to normal tissue-matched samples-most notably with breast cancer [60], melanoma [61] and germ cell tumors [62]. More recent preliminary studies in hepatocellular carcinoma [63] and pancreatic adenocarcinoma [64] have also found increased HERV-K mRNA levels in cancer as compared to normal tissues. In all of these tumors, significant debate remains as to whether HERV-K expression acts as a disease driver or is simply a result of global epigenetic changes in the cancer cell [65]. Though causality remains unclear, there is clear evidence that HERV-K expression can have significant effects on the transcriptome of the cancer cell through insertional position near, for example, proto-oncogenes, or through acting as alternative promoters, polyadenylation signals or alternative splice sites within introns of human genes [66-68]. HERV-K-*env* expression has specifically been linked to perturbations in both the p53 and RAS signaling pathways in breast cancer [69], while in pancreatic adenocarcinoma, RNA-seq analysis following sh-RNA knock-down of HERV-K-*env* showed decreased expression of genes on the RAS-ERK-RSK pathway [64].

Equally important, HERV-K expression has been proposed as an important potential biomarkers for all the above diseases [15, 16], due in part to the very low expression levels in corresponding normal tissue. In breast cancer, HERV-K expression was found to effectively differentiate basal cell carcinoma from other breast cancer subtypes [34]; a clinically important finding given the aggressive nature of this tumor. Like many pediatric solid organ tumors, the next steps in hepatoblastoma therapy involves finding appropriate molecular targets for immunotherapy, as well as the ability to molecularly stratify clinically aggressive tumors. This has led multiple groups to explore the potential role of both microRNAs and long non-coding RNA (lncRNAs) in hepatoblastoma [70-72]. However, the field remains limited by a lack of preserved tissue, including normal patient-matched tissue controls that would enable a more extensive molecular analysis of these rare but important childhood tumors.

### 4.2 Importance of RNA-seq Analysis Pipelines for HERV-K Expression and its Limitations

With the advancement in massive parallel sequencing technologies, which have now generated numerous RNA-seq datasets of multiple tumor types, bioinformatic techniques that allow analysis of these powerful datasets are becoming increasingly necessary [73]. HERV-K proviruses pose a difficult challenge for computational analysis of RNA-seq data as these regions are poorly annotated and constitute repeat elements making unique alignment of RNA-seq reads difficult [49]. As a result of these limitations, the vast majority of investigations into HERV-K RNA expression in cancer have not been able to localize expressed RNAs to individual HERV-K proviral loci. This has fundamentally limited the ability to discover how these proviruses may affect cell biology and identify potential translated HERV-K proteins that may also affect cell biology, as well as present unique targets for immunotherapy. Nevertheless, given the abundance of HERV sequences in the human genome, a complete understanding of human cancer biology will undoubtedly require elucidating the effects that these entities have on the cellular machinery.

The methodology presented in this investigation allows for simple analysis and screening of RNA-seq datasets for HERV-K expression. Furthermore, methods used to quantify read counts across specific proviral loci allowed for a fuller description of the HERV-K expression, in addition to simple detection of the presence or absence of HERV-K mRNA. In our analysis, we used the Salmon alignment tool in transcriptome alignment mode (pseudoalignment). As Salmon identifies unique k-mers across the transcriptome, we were thus able to effectively distinguish unique k-mers across the different HERV-Ks and could assign uniquely aligned reads to individual proviruses. The alignment tool then allocated multi-map reads to proviral locations that had at least one unique alignment across the provirus. The limitation of the Salmon pseudoalignment analysis was that we could not determine where reads were aligning across the provirus. As the proviruses themselves have complex transcriptomes, some with large insertions or deletions of nucleotides (i.e. indels), this was necessary to infer the potential proviral mRNAs and proteins that could be produced. The use of a standard RNA-seq alignment tool HiSAT2 accomplished this task, though we still had to control for multi-mapped reads which was accomplished with Samtools. With this step, we lost ∼20-25% of the total reads that aligned to HERV-K loci. While this was acceptable for mapping reads across different regions of the proviruses, it confounded effective quantification. Furthermore, multi-map reads are more likely to occur with more recently integrated proviruses that are more complete [49]. This is unfortunate as these proviruses are also the most likely to produce viral proteins and therefore have the largest potential effect on the cell.

### 4.3 Overall Limitations of this Study

There are several limitations in the dataset that we used in this analysis. The use of non-patient matched liver control tissue prevented a more thorough analysis of differential expression between tumors and normal liver, given that HERV-Ks remain polymorphic in the human population. A lack of aged-matched and patient-matched normal tissue controls is a common issue with current RNA-seq analysis studies of fetal solid organ malignancies. Furthermore, the small dataset did not allow the ability to correlate HERV-K transcription with clinical outcomes, including stage of disease, reoccurrence, disease resistance and over-all survival. It also limited the potential to correlate HERV-K expression with differentiation/histologic subtypes of hepatoblastoma including fetal, embryonal and undifferentiated disease. Lastly, from a bioinformatics perspective, a future annotation of all viral mRNAs-including Gag, Env, Rec and/or Np9 that can be produced from each individual provirus, will allow for direct screening of viral mRNAs that have the potential to encode for viral proteins.

## 5. Conclusion

The current investigation demonstrates that several Human Endogenous Retrovirus-K proviruses are transcribed in hepatoblastoma, with increased RNA expression from several proviral loci in hepatoblastoma as compared to normal liver controls. The large difference in HERV-K expression profiles between hepatoblastoma and normal liver sheds light on several important questions that make HERV-K studies important. Future investigations are required both to explore HERV-K expression as a tool for molecular disease stratification, as well as for targeted immunotherapy. This study also highlighted the complex nature of HERV-K RNA expression in hepatoblastoma. Hepatoblastoma samples demonstrated large differences in overall expression profiles of HERV-K, both in total aligned reads to HERV-K proviruses and in the total number of expressed proviruses. Finally, our study highlights the important need to continue to develop tumor banks for pediatric solid organ tumors that include patient matched tissue controls for appropriate molecular comparison.

## Data Availability

Data is publicly available.

## Acknowledgements

The authors kindly acknowledge Dr. Rakesh Sindhi of UPMC, who made the Hepatoblastoma RNA-seq dataset available for this analysis. Partial salary support for MLH and DR was provided by the Charles H. Ross Jr and Myles H. Thaler Professorship endowments at the University of Virginia. This work was supported by The National Cancer Institute of the National Institutes of Health (Grant numbers: T32 CA163177 and R01 CA206275).

## Abbreviations

(HERV-K): Human Endogenous Retrovirus-K
(HB): Hepatoblastoma
(FT): Fetal Tumor
(NC): Normal Control

---

Supplemental File 2- Significant Differential Gene Expression Analysis (Pseudoalignment with Salmon, Differential Gene Expression with DESeq2) of High HERV-K Expression Hepatoblastoma Samples compared to Low HERV-K Expression Hepatoblastoma Samples (top 100 genes ranked by log2fc).

| Gene_Symbol | Gene_Ensembl_ID | baseMean | log2FoldChange | padj |
| --- | --- | --- | --- | --- |
| AC010970.1 | ENSG00000225840 | 4091871.811 | 25.58649925 | 5.22E-17 |
| RTL1 | ENSG00000254656 | 33.05735061 | 23.55870912 | 1.27E-11 |
| RNA5SP202 | ENSG00000201185 | 1083.282101 | 14.68388487 | 3.11E-05 |
| NOS1AP | ENSG00000198929 | 106.6727031 | 11.33920845 | 0.002516312 |
| ZACN | ENSG00000186919 | 92.83449212 | 11.14137145 | 0.001725542 |
| LRRC75A | ENSG00000181350 | 229.2868567 | 10.92063082 | 0.000719551 |
| LRRC37A16P | ENSG00000267023 | 106.1675703 | 10.89945798 | 0.001503372 |
| RNA5SP149 | ENSG00000201822 | 65.61712433 | 10.63275295 | 0.007560641 |
| ERICH3 | ENSG00000178965 | 64.63895786 | 10.59237992 | 0.017724784 |
| RAB11FIP1P1 | ENSG00000228492 | 61.1815703 | 10.5316707 | 0.008119703 |
| TMEM52B | ENSG00000165685 | 57.49828505 | 10.45803747 | 0.005406543 |
| CICP14 | ENSG00000281490 | 57.28417004 | 10.4260215 | 0.009696774 |
| AC010507.1 | ENSG00000282051 | 56.42677775 | 10.40690811 | 0.009696774 |
| SPDYE10P | ENSG00000274570 | 56.2187497 | 10.40622302 | 0.004042745 |
| SPDYE3 | ENSG00000214300 | 50.87409277 | 10.25251506 | 0.01095637 |
| SPDYE2 | ENSG00000205238 | 44.04967381 | 10.03982686 | 0.012274692 |
| SPDYE1 | ENSG00000136206 | 43.6652868 | 10.02878074 | 0.011793257 |
| CEMP1 | ENSG00000205923 | 39.301951 | 9.880801846 | 0.012059115 |
| AL590627.1 | ENSG00000255585 | 36.28950676 | 9.767233131 | 0.012634784 |
| AC098934.1 | ENSG00000214796 | 34.98115507 | 9.739083023 | 0.004211152 |
| ANKRD19P | ENSG00000187984 | 82.26444706 | 9.618303746 | 0.004805867 |
| AC006116.7 | ENSG00000267459 | 31.97610894 | 9.571424785 | 0.016993644 |
| HIST2H2AC | ENSG00000184260 | 30.99940737 | 9.552938905 | 0.017073074 |
| SPDYE15P | ENSG00000278080 | 28.42695147 | 9.407594267 | 0.017147805 |
| OR2A7 | ENSG00000243896 | 28.27954406 | 9.383537617 | 0.034217837 |
| AC239799.1 | ENSG00000213240 | 26.70009139 | 9.315613897 | 0.018440141 |
| RIMKLBP1 | ENSG00000189089 | 26.25957663 | 9.274331333 | 0.036329659 |
| AC018638.1 | ENSG00000229413 | 25.76126588 | 9.264153505 | 0.01938907 |
| NAP1L4P1 | ENSG00000177173 | 25.733931 | 9.261757469 | 0.019115839 |
| NA | ENSG00000283186 | 84.03591027 | 9.225414142 | 0.005635334 |
| UHRF2P1 | ENSG00000223650 | 24.66585027 | 9.200871906 | 0.020150585 |
| SLC5A2 | ENSG00000140675 | 147.0998851 | 9.18225622 | 0.00379274 |
| TNFSF4 | ENSG00000117586 | 40.11592425 | 9.152166319 | 0.002106795 |
| AC138409.1 | ENSG00000215156 | 76.83588876 | 9.145051041 | 0.009696774 |
| AL671277.1 | ENSG00000227766 | 23.02087131 | 9.124723718 | 0.020019583 |
| ABCC11 | ENSG00000121270 | 23.07506169 | 9.095204882 | 0.02225255 |
| EPS8L3 | ENSG00000198758 | 68.88099066 | 9.086998608 | 0.008139284 |
| LHX4 | ENSG00000121454 | 22.2219767 | 9.083985193 | 0.011513186 |
| NSRP1P1 | ENSG00000235613 | 22.64124858 | 9.077155283 | 0.021793467 |
| RPL6P2 | ENSG00000227056 | 22.21964176 | 9.027468219 | 0.042060283 |
| CLDN22 | ENSG00000177300 | 21.02849797 | 8.959460497 | 0.024393675 |
| GPRC5D | ENSG00000111291 | 20.17251593 | 8.915335381 | 0.024451427 |
| SKINT1L | ENSG00000242267 | 101.0365844 | 8.891302102 | 0.009696774 |
| SATL1 | ENSG00000184788 | 18.80398912 | 8.788277967 | 0.028224944 |
| CCDC114 | ENSG00000105479 | 19.94194504 | 8.746077984 | 0.018114198 |
| AC097063.1 | ENSG00000224570 | 18.01486771 | 8.722519461 | 0.030258333 |
| CDR1 | ENSG00000184258 | 17.24731336 | 8.709381632 | 0.047699515 |
| NUF2 | ENSG00000143228 | 16.51864422 | 8.687687009 | 0.018181527 |
| AC112497.1 | ENSG00000279413 | 17.12815226 | 8.685214698 | 0.029019188 |
| AC138951.1 | ENSG00000233974 | 17.24677605 | 8.678731827 | 0.028695264 |
| ALOX12B | ENSG00000179477 | 17.0695303 | 8.678484582 | 0.033061621 |
| AL627230.7 | ENSG00000278134 | 16.40468653 | 8.634912255 | 0.033393099 |
| PIK3C2G | ENSG00000139144 | 150.2972016 | 8.629295101 | 0.024739607 |
| TRIM67 | ENSG00000119283 | 16.66747848 | 8.628735318 | 0.030198847 |
| ACAN | ENSG00000157766 | 22.44510057 | 8.619269945 | 0.014164622 |
| IPP | ENSG00000197429 | 29.47080519 | 8.601740121 | 0.004211152 |
| MMP25 | ENSG00000008516 | 17.55215 | 8.595807633 | 0.019115839 |
| POTEI | ENSG00000196834 | 15.7348339 | 8.576470526 | 0.034687012 |
| TMEM262 | ENSG00000187066 | 15.98396235 | 8.57244938 | 0.031108731 |
| CR788268.1 | ENSG00000270909 | 15.70310704 | 8.564887943 | 0.032799305 |
| DNAH11 | ENSG00000105877 | 16.02935806 | 8.561856152 | 0.032052746 |
| AL589880.1 | ENSG00000229939 | 24.04372433 | 8.552978051 | 0.012059115 |
| NRIP2 | ENSG00000053702 | 15.55135525 | 8.549996031 | 0.020478645 |
| SYCP3 | ENSG00000139351 | 15.80577069 | 8.545541485 | 0.032052746 |
| NA | ENSG00000233438 | 15.73030109 | 8.530475088 | 0.016228556 |
| CXCR5 | ENSG00000160683 | 25.881473 | 8.499874845 | 0.014261537 |
| DUSP28 | ENSG00000188542 | 61.70692827 | 8.493280426 | 0.00088236 |
| KAAG1 | ENSG00000146049 | 14.86274184 | 8.483865475 | 0.016180782 |
| ODCP | ENSG00000244556 | 15.01245265 | 8.467797921 | 0.034177988 |
| PRR27 | ENSG00000187533 | 16.03140675 | 8.459909182 | 0.036078213 |
| ZBTB45P1 | ENSG00000225108 | 14.7456211 | 8.42963194 | 0.036141391 |
| KCNK12 | ENSG00000184261 | 14.53838021 | 8.428479057 | 0.03487786 |
| C6orf201 | ENSG00000185689 | 14.32142725 | 8.39426497 | 0.036141391 |
| JSRP1 | ENSG00000167476 | 32.27298907 | 8.377024906 | 0.012634784 |
| FKBP6 | ENSG00000077800 | 14.07194039 | 8.372071538 | 0.036639762 |
| AL583844.1 | ENSG00000227236 | 13.73637294 | 8.347469364 | 0.018279268 |
| AD000091.1 | ENSG00000267135 | 202.1551464 | 8.32351416 | 0.006082821 |
| CFAP73 | ENSG00000186710 | 13.58842914 | 8.300835516 | 0.028964335 |
| IL15 | ENSG00000164136 | 33.87278437 | 8.278196009 | 0.013892554 |
| CHRM5 | ENSG00000184984 | 13.06858868 | 8.265718545 | 0.039344501 |
| DLEC1 | ENSG00000008226 | 80.37672972 | 8.260461406 | 0.015205586 |
| HEPN1 | ENSG00000221932 | 34.21641499 | 8.229411046 | 0.029969725 |
| TATDN2P2 | ENSG00000218226 | 12.5365546 | 8.217029887 | 0.041350728 |
| NUDT4P2 | ENSG00000271121 | 12.23890901 | 8.196089684 | 0.04262468 |
| LRRC46 | ENSG00000141294 | 12.23589239 | 8.17904649 | 0.042104363 |
| AC012513.2 | ENSG00000236478 | 12.26242788 | 8.16948291 | 0.04262468 |
| GPR39 | ENSG00000183840 | 20.26106046 | 8.131776218 | 0.020563205 |
| SRRM5 | ENSG00000226763 | 11.84078726 | 8.12465022 | 0.044146341 |
| RPL7P9 | ENSG00000137970 | 52.31788937 | 8.124477932 | 0.026486379 |
| SMARCB1 | ENSG00000099956 | 29.35146147 | 8.107417268 | 0.012069371 |
| SPRN | ENSG00000203772 | 11.04191731 | 8.082624987 | 0.047977544 |
| DDX3P1 | ENSG00000232928 | 11.29124671 | 8.075265182 | 0.046639931 |
| FAM216A | ENSG00000204856 | 47.59781672 | 8.051949587 | 0.00887882 |
| RAD51AP1 | ENSG00000111247 | 11.246235 | 8.035914549 | 0.029969725 |
| IQCC | ENSG00000160051 | 19.85514963 | 8.032039212 | 0.011269771 |
| HIST2H4A | ENSG00000270882 | 10.64297921 | 7.998016381 | 0.021046164 |
| RNF32 | ENSG00000105982 | 25.7994181 | 7.958537953 | 0.027169179 |
| SLC30A3 | ENSG00000115194 | 9.707443186 | 7.957551833 | 0.038639101 |
| CCDC39 | ENSG00000145075 | 12.59878428 | 7.941085757 | 0.029969725 |
| TDRD6 | ENSG00000180113 | 9.751585097 | 7.930051589 | 0.024981655 |

Supplemental Figure 3. Gene Ontology (GO) terms for cellular localization (Supplemental Table 1) and molecular function (Supplemental Table 2) as well as enriched Kyoto Encyclopedia Genes and Genomes (KEGG) (Table 3) terms for significantly differentially expressed genes between high HERV-K expressing hepatoblastoma samples and low HERV-K expressing hepatoblastoma samples.

**Supplemental Table 1:**
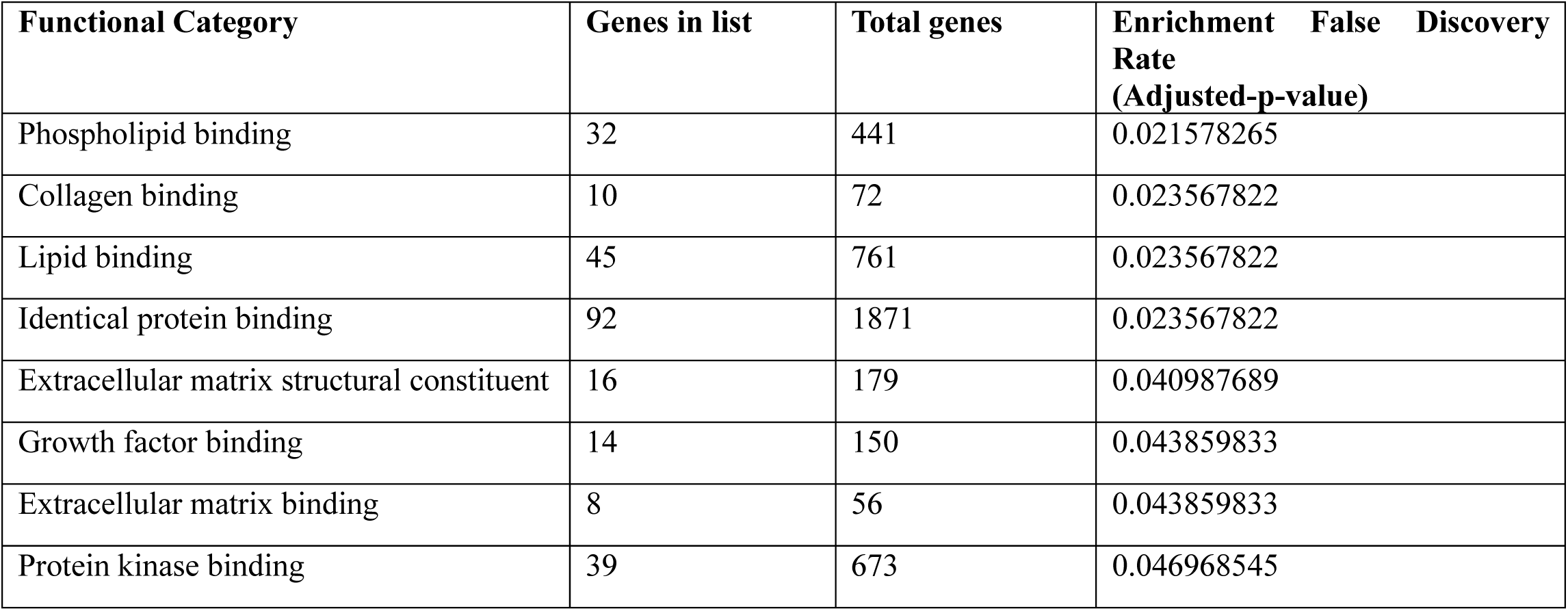
Gene Ontology (GO) molecular function analysis following differential gene expression analysis of high HERV-K expressing Hepatoblastoma vs low HERVK expressing Hepatoblastoma

**Supplemental Table 2:**
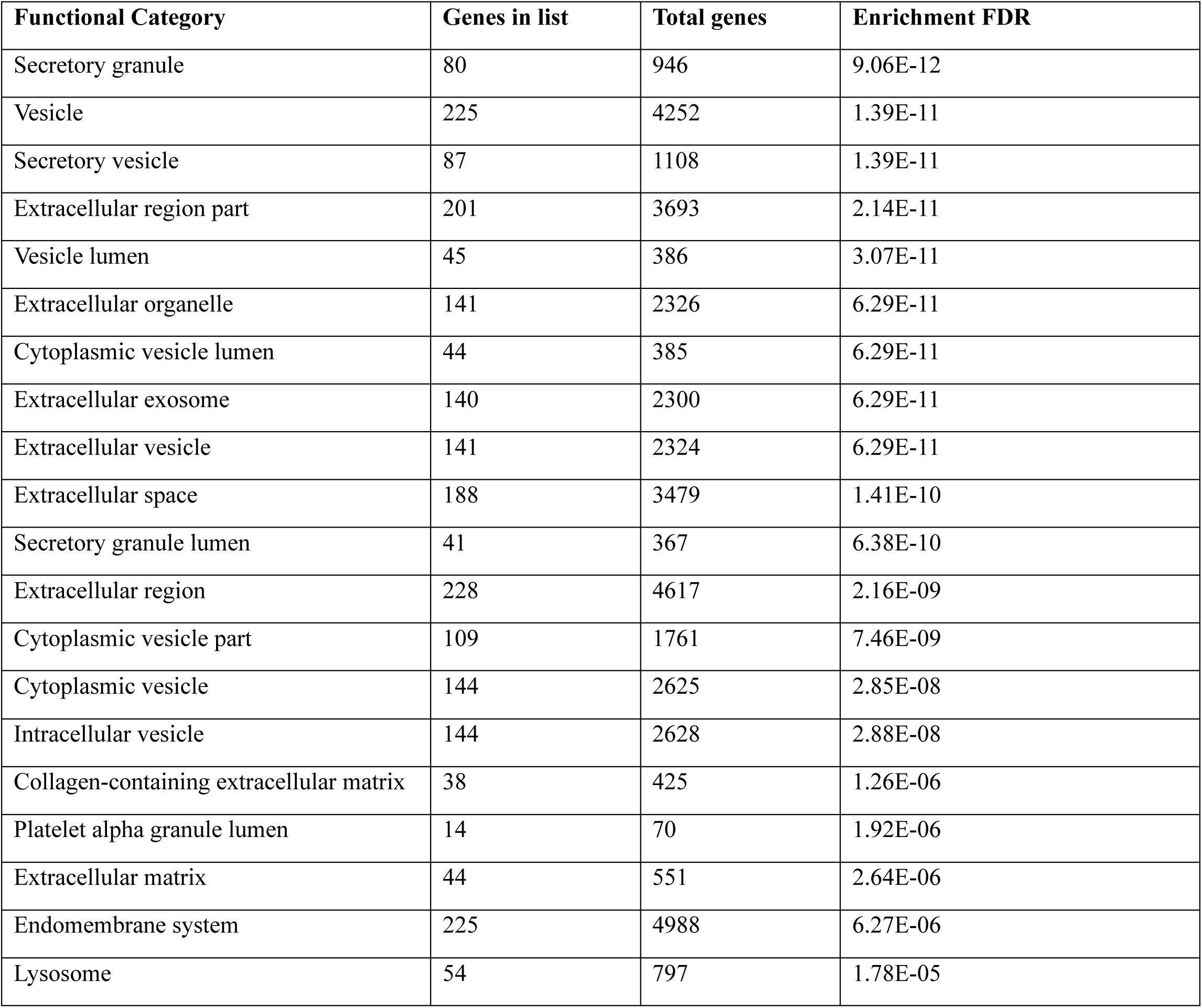
Gene Ontology (GO) cellular localization analysis following differential gene expression analysis of high HERV-K expressing Hepatoblastoma vs low HERVK expressing Hepatoblastoma (Top 20 terms)

**Supplemental Table 3:**
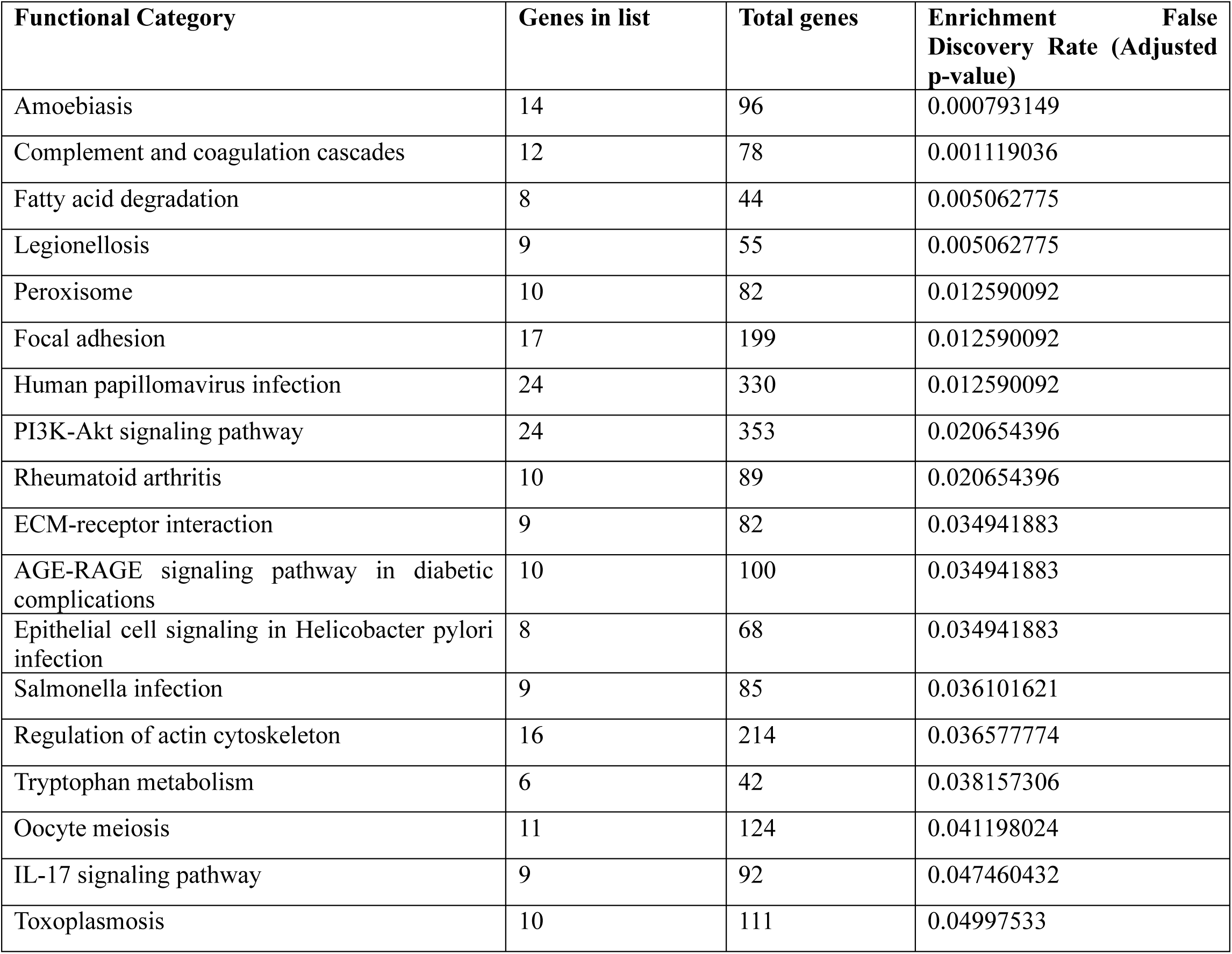
Kyoto Encyclopedia of Genes and Genomes Enrichment Analysis following differential gene expression analysis of high HERV-K expressing Hepatoblastoma vs low HERVK expressing Hepatoblastoma.

